# Stakeholder perspectives on the barriers and facilitators to integrating cardiovascular disease and diabetes management at primary care in Kenya

**DOI:** 10.1101/2024.12.25.24319646

**Authors:** Elvis O. A. Wambiya, James Odhiambo Oguta, Robert Akparibo, Duncan Gillespie, Peter Otieno, Catherine Akoth, Jemima Kamano, Peter Kibe, Yvette Kisaka, Elizabeth Onyango, Gladwell Gathecha, Peter J. Dodd

## Abstract

Integrated care is increasingly recognised as a key strategy for the management of multimorbidity. However, evidence on the factors associated with the implementation of integrated care models in low- and middle-income countries remains limited. We explored how stakeholders perceived integrated care, existing challenges, and recommendations for integrating cardiovascular disease and diabetes management at primary health care (PHC) level in Kenya. We conducted a qualitative study using key informant interviews with 16 key stakeholders involved in healthcare delivery, research, and policy on non-communicable diseases (NCDs) in Kenya between February and March 2024. All interviews were audio recorded and transcribed; and the data analysed both inductively and deductively within NVivo software. The deductive analysis was guided by the Rainbow Model of Integrated Care (RMIC) framework, which combines key dimensions necessary for successful integrated care with key elements of primary care. The RMIC framework dimensions include systems, clinical, organisational, professional, functional, and normative integration. Out of the six RMIC dimensions, stakeholders’ description of integrated care included elements of clinical, systems, and professional integration. Key systems level challenges included disparity between policy and practice, inadequate resource allocation, donor-driven priorities, and limited stakeholder collaboration. Fragmented care delivery was a key organisational challenge. Limited resources for integrated care delivery and inadequate staff numbers and capacity were considered key challenges in the functional and professional dimensions of the RMIC framework. Additional challenges included ‘siloed’ mindset (normative) and limited evidence on effective or cost-effective integrated care models. To address the identified barriers, policy-makers should develop clear and adaptable how-to county-specific guidelines for implementation and evaluation of integrated care at PHC level. There is a need for advocacy and research on models of integrated care at PHC level to guide prioritization and resource allocation in Kenya.

## Introduction

Non-communicable diseases (NCDs) cause a substantial disease burden globally fuelled by changing social, economic, and structural factors [1]. NCDs account for 74% of mortality worldwide, and 77% of mortality in low-and middle-income countries (LMICs) [2].

Cardiometabolic diseases, including cardiovascular diseases (CVDs) and diabetes, are among the leading causes of NCDs morbidity and mortality globally [3–6]. The global increase in NCDs has contributed to the growing number of people living with multimorbidity, the co-existence of two or more chronic diseases in an individual [7–9]. In addition to increased disability and reduced quality of life, multimorbidity increases financial burden for patients navigating care and places greater demands on health systems [10–14]. Because of shared pathophysiology and risk factors, multimorbidity involving cardiometabolic diseases such as hypertension and diabetes is common [15–17].

Integrated care, which reduces fragmentation and promotes comprehensive and coordinated health service delivery, is increasingly recognised as a key strategy for the management of multimorbidity [18–21]. While more evidence on integrated care exists from high-income countries (HICs), LMICs including sub-Saharan Africa (SSA) are gradually adopting integrated care models, some of which have shown promise in improving outcomes for people with multimorbidity [22–28]. However, evidence on the factors associated with the implementation of integrated care models in LMICs remains limited.

While several integrated care frameworks such as the chronic care model [29,30], the WHO framework on integrated people-centred health services [31], and the rainbow model of integrated care (RMIC) [32] have been employed in HICs to assess and guide integrated care implementation, their application and adaptation in LMICs are underexplored. The RMIC posited by Valentijn *et al.* (2013), provides a comprehensive framework to understand integrated care by combining primary health care functions with six interrelated dimensions of integration namely: clinical, professional, organisational, systems, functional, and normative integration [32]. By encompassing both micro-level care processes and macro-level health system structures, the RMIC allows for a nuanced understanding of barriers and facilitators across diverse levels of the health system. The RMIC has been employed in integrated care studies in HICs [33,34] and LMICs [35–38].

In Kenya, NCDs account for 41% of all deaths annually and have resulted in significant economic challenges, including reducing household income by more than 28% [39]. Notably, CVDs and diabetes are among the leading causes of the burden of NCDs in Kenya, accounting for 36% and 6% of total NCDs deaths in 2019 respectively [39]. Consequently, CVDs and diabetes comorbidity is increasingly becoming a concern in Kenya [40–42]. Primary health care (PHC) is central to addressing the dual burden of communicable and non-communicable diseases in LMICs including Kenya. Strengthening PHC by integrating NCD prevention and management is critical to improving health outcomes, reducing economic losses, and achieving universal health coverage (UHC).

Despite the inclusion of integrated care in existing policies and guidelines in Kenya including the Kenya Health Policy and National NCD Strategic Plan [39], evidence on integration of NCDs at primary care level remains scarce. Understanding the barriers, facilitators, and best practices for implementing integrated care models is critical to addressing the challenge of multimorbidity in Kenya. This RMIC framework was considered particularly suitable for this study as it offers a multidimensional lens to contextually explore how prevention and management of NCDs can be effectively integrated at the PHC level in Kenya. Guided by the RMIC framework, this study explored stakeholders’ contextual definition of integrated care, challenges for implementation, and recommendations for effective integration of CVDs and diabetes management at PHC level to tackle the emerging challenge of cardiometabolic multimorbidity in Kenya.

## Methods

### Study design and setting

This was a qualitative study that employed key informant interviews (KIIs) to explore stakeholder perceptions on their contextual definition, challenges, and recommendations for implementing integrated care at primary care level in Kenya. Kenya, a country in Eastern Africa, is classified as a lower-middle-income economy [43]. Under its new constitution enacted in 2010, healthcare became a devolved function in Kenya whereby health service delivery is a function of the 47 semi-autonomous county governments with the national government retaining a policy and regulatory role [44]. This study was designed and conducted according to the Standards for Reporting Qualitative Research (SRQR) [45].

### Participants

Participants were key stakeholders involved in health care delivery, health policy, research, and decision-making for NCDs in Kenya. These included policy or decision-makers and technical personnel at the ministry of health, researchers from government parastatals and non-governmental organisations (NGOs), academics, and health economists involved in economic evaluations of interventions for chronic diseases.

### Sampling and recruitment

The stakeholders were selected using purposive sampling based on their involvement in chronic disease/ NCD prevention and control in Kenya as a policy or decision-maker, technical personnel, researcher, health care provider, or academic. A master list of key stakeholders was developed from a document review conducted prior to the study and shared with a key contact from the Ministry of Health. The key contact supplemented the list of stakeholders based on their knowledge and engagements with other similar stakeholders. Recruitment was conducted between 05 October 2023 and 30 January 2024.

### Data collection

Data were collected from the key informants using a key informant interview guide between 01 February 2024 and 02 March 2024. The lead author (EW) led the interviews and was assisted by one of the co-authors (JO). The interview guide had three main parts: (1) stakeholders’ contextual understanding of integrated care, (2) challenges of effectively integrating chronic NCD care at PHC level, and (3) recommendations to address any perceived barriers to implementing integrated care at PHC level to address cardiometabolic multimorbidity (Supplementary note 1). We defined cardiometabolic multimorbidity as the existence of two or more chronic diseases in the same individual, at least one of which was a cardiometabolic disease. To align the definition to the Kenyan context, we focused on CVD and diabetes comorbidity based on the high burden and asked the stakeholders to picture integration at the PHC level in the context of these diseases and similar conditions. The interviews were conducted in a private and quiet location convenient to the participants. The interview guide was pilot tested with two participants who did not form part of the final sample prior to actual data collection. All interviews were conducted in English language and were approximately an hour long. Each interview was audio recorded following written informed consent. The researchers continued enrolling stakeholders for interviews until data saturation was achieved.

### Conceptual framework

We used the RMIC by Valentijn *et al.*, (2013) (Figure 1) to guide the design, data collection and analysis. This model highlights six key dimensions crucial for successful integrated care (clinical, professional, organisational, system, functional, and normative) structured around the macro, meso, and micro levels of the health care system, combined with the integrative functions of primary care. The definitions of framework dimensions are shown in Supplementary table 1.

**Figure 1:**
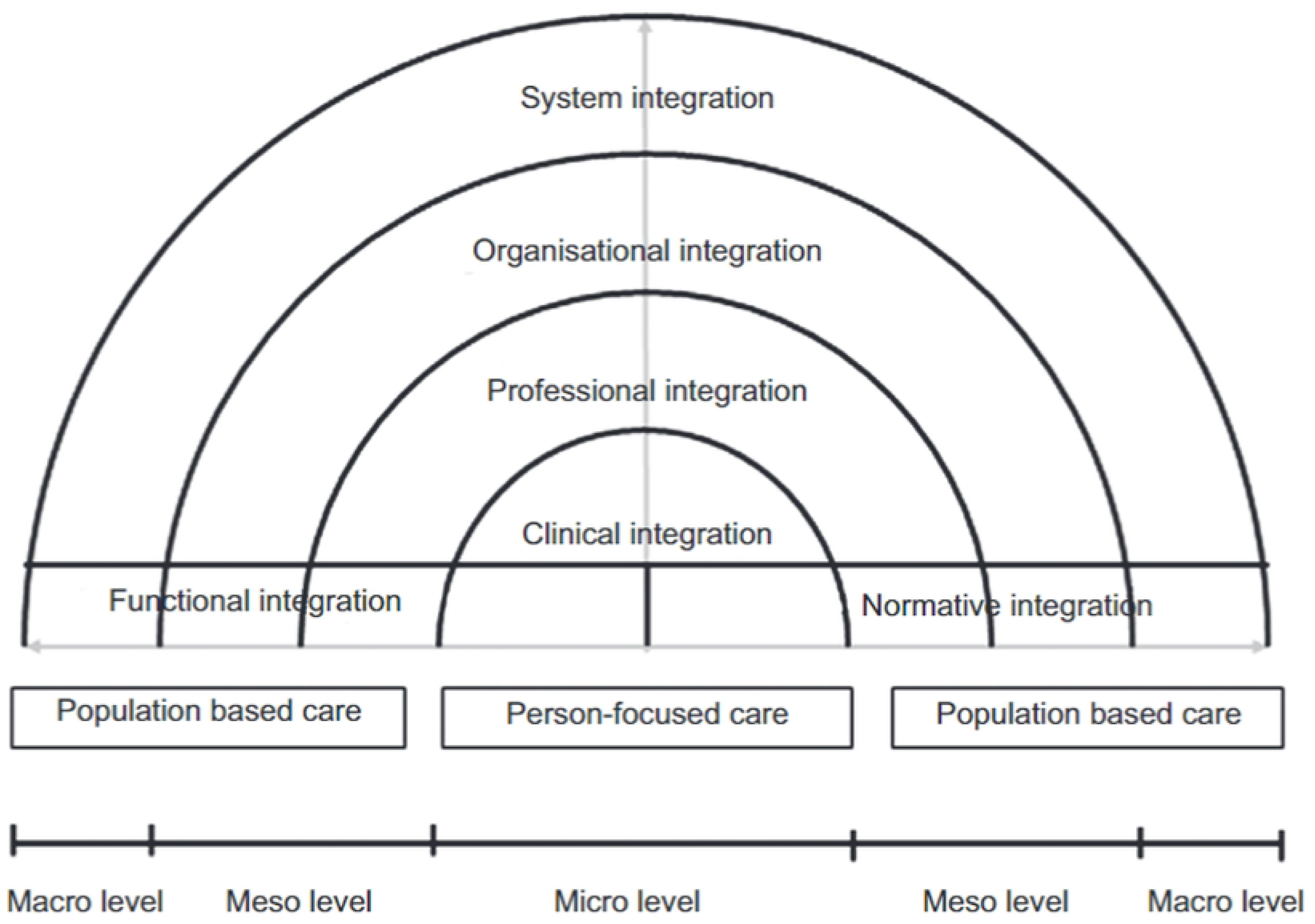
Rainbow Model of Integrated care conceptual framework. Adapted from Valentijn *et al.,* (2013)

### Data analysis

Thematic analysis involving a combination of inductive and deductive methods, guided by the RMIC [46] (Figure 1), was used. Data familiarisation through immersion into the study transcripts was conducted [47], after which a codebook was developed based on the RMIC framework dimensions (Figure 1). Qualitative data analysis and management software - NVivo version 14 was used to systematically code relevant data segments across transcripts into main themes and sub-themes based on the code book. Two researchers (EW and JO) coded the first five transcripts, which were cross-checked by RA and CA. One researcher (EW) then used the validated codebook to code the remaining transcripts. While text segments from the transcripts that matched the predefined codes were added against them, new emerging themes and sub-themes that did not fit into the framework were developed and added as new themes and sub-themes. Finally, the identified themes were integrated and synthesised for interpretation and reporting [48].

### Ethical approval

The study was approved by Moi Teaching and Referral Hospital/ Moi University Institutional Research and Ethics Committee (approval number 0004625), the University of Sheffield SCHARR research ethics committee, and a research permit granted by the National Commission for Science Technology and Innovation (NACOSTI) in Kenya (License number: NACOSTI/P/23/31904). All participants were taken through a detailed information sheet about the study after which they provided written informed consent prior to participation in the study. All the transcripts were anonymised during analysis and reporting to ensure confidentiality of study participants. The data were stored in a secure University of Sheffield file store only accessible to the researcher and the study team.

## Results

### Participant characteristics

Overall, 16 stakeholders were interviewed (Table 1), the majority of whom were females. Three stakeholders who had initially agreed to the interviews could not participate due to unforeseen circumstances. In terms of their professional background, the majority of the participants referred to themselves as public health specialists with a clinical background (n=9). Seven of the 16 participants were affiliated with the government at either national, county level, or through a parastatal. Six of the 16 participants were involved in policy making for NCDs. On average, participants had 18 years (Mean = 18.2, SD = 8.6) of professional experience, about 10 years (Mean = 9.7, SD = 6.1) of which were specifically related to NCDs.

**Table 1:** Characteristics of the study participants.

| **Participant characteristic** | **Frequency** |
| --- | --- |
| Sex |  |
| Female | 10 |
| Male | 6 |
| Affiliation |  |
| National parastatal | 1 |
| County health department | 2 |
| Ministry of Health (National level) | 4 |
| Non-governmental organisation | 7 |
| University | 2 |
| Occupation |  |
| Academic | 2 |
| Clinician | 1 |
| Health economist | 1 |
| MD + Public Health specialist | 9 |
| Pharmacist + Public Health specialist | 1 |
| Policy advocate | 1 |
| Public Health specialist | 1 |
| Involvement in policy making |  |
| Yes | 6 |
| No | 10 |
| **Total** | **16** |

### Stakeholders understanding of integrated care

The description of integrated care in the Kenyan context according to the stakeholders was diverse and mainly fell within the clinical, systems and professional integration dimensions of the integrated care framework.

### Clinical integration

Most stakeholders perceived integrated care to be the provision of holistic and patient-centred care in a “one stop shop”. In their view, this meant the provision of prevention and treatment health services for different chronic diseases in the same facility. They considered it to be care that considers the patient as a whole and looks beyond the disease. The below quotes are from three stakeholders giving their remarks:

> “If you go to a health facility right now for example cervical cancer is screened at MCH, hypertensives are mainly seen at medical outpatient clinics, people with diabetes are seen at diabetes clinic…but for me integration would mean we have that one stop where any matters NCDs are seen at a single stop or clinic or something of sort.” (Female respondent, policy maker, T1)

> “The ideal would be one chronic disease clinic where everybody there can be managed, and all their conditions are addressed in one stop shop and then you don’t have to have these back and forth.” (Male respondent, clinician, and public health specialist, T16)

> “Also, integration is trying to synergize resources, trying to see how as the same health worker I can see both diabetes and hypertension then how do we facilitate that?

So, trying to maximise resources by seeing how they can serve different purposes. The same resource can be used to address different conditions.” (Female respondent, clinician and public health specialist, T11)

### Systems integration

At the systems level, integrated care was perceived by half of the respondents as that which was not vertical or fragmented as has been observed in various funded projects such as HIV or NCD-specific studies. Four participants involved in policy making also perceived that integration starts with the development of guidelines, protocols, and standards that promote the collaborative and coordinated delivery of care by health personnel. This was exemplified through the following quotes:

> “Stop the silo mentality that someone is treating X, or my grant is only supposed to facilitate this. As you know previously, HIV funding was just for HIV and even though it made a lot of sense you try and integrate things into that. There were clear directions that you cannot do that.” (Female respondent, public health specialist, T10)

> “It really involves developing new protocols that involve the physicians or the service providers …working together in a multidisciplinary team…so there is a need to build those multidisciplinary teams to re-evaluate, re-examine how best to deliver that to re-engineer our system…” (Female respondent, clinician and public health specialist, T14)

### Professional integration

Linked to clinical integration, nine stakeholders who had been involved in clinical practice during their careers perceived that integrated care involved the interdisciplinary coordination by the healthcare professionals to deliver comprehensive health services to patients with multiple diseases. This is highlighted in the following selected quotes:

> **“**So, my ideal integration is that the patient who has more than one condition, when they come to the same clinic, they should be served…if the specialists are different at least their clinics should be on the same day. There should be a multimorbidity clinic where there is a specialist who can attend to most of the things.” (Male respondent, clinician and public health specialist, T15)

> “It really involves now developing new protocols that involves the physicians or the service providers to be able to attend to this particular person, but you know working together a multidisciplinary team is actually attending to this particular patient at one point in time” (Female respondent, clinician and public health specialist, T14)

### Challenges of implementing integrated care for cardiometabolic multimorbidity in Kenya

#### Systems level

##### Disparity between policy and practice

The disparity between policy and practice in the implementation of integrated care in Kenya was evident from the interviews. Despite having well-developed policies and frameworks in place for NCDs prevention and control, there is a significant gap in actual implementation. During the interviews, stakeholders attributed this gap to a lack of resources, including personnel, coordination, and financial support. Stakeholders pointed out that while policy documents exist, the challenge lies in translating these policies into actionable steps and ensuring that the intended services are effectively delivered at all levels. This was summarised by two stakeholders who remarked that:

> “…This is where I would say the policy is lacking, the policies are there, they are laid down but implementation is lacking in terms of resources, be it personnel, be it coordination, be it finances, I think that is where we are lacking in terms of improving non-communicable diseases control…The policy makers have the documents but are we doing what is laid down? I think that is lacking.” (Female respondent, University lecturer and public health specialist, T9)

> “To be able to offer these services at that level then has not been substantially done, and I think it’s a hole. It’s a gap in translating policy to implementation. And maybe that’s also another policy conversation. Like how do we translate policy to implementation?” (Male respondent, clinician and public health specialist, T16)

##### Prioritisation of integrated care

Some stakeholders felt that integrated care or multimorbidity prevention and control have not been prioritised in health care policy and programs in Kenya. This was evident from the following two quotes from the participants:

> “I would say it hasn’t been prioritised…although it is in NCDs policy documents as a collection of conditions, but the concept of multimorbidity hasn’t been highlighted as much as it should, I would say.” (Male respondent, clinician and health economist, T5)

> “So, it’s something that we need to focus on in terms of integrating other conditions and multimorbidity and so on. I don’t think there has been a big emphasis on it at the moment, but it is something that we definitely need to focus on.” (Female respondent, public health specialist, T10)

##### Lack of clear policies and guidelines on integrated care or multimorbidity

It also emerged from the participants that while strategic plans include components of integration, there is no direct, comprehensive policy explicitly addressing the integration of NCDs with other conditions. Stakeholders indicated that existing policies, such as task sharing, have faced resistance and lacked clear guidance, causing confusion and hindering effective integration. They also highlighted the lack of specific guidelines addressing multimorbidity, with existing guidelines focused on individual diseases like hypertension and diabetes. This is exemplified in the following quotes:

> “I can’t say that we have a direct policy on integration, but if you look at our strategic plan it has some component which is talking of integration, a very huge component of integrating NCDs with other conditions. So, we don’t have that direct document but within our strategic plan we have a whole component on integration.” (Male respondent, clinician and public health specialist, T13)

> “I know there was a task sharing policy, but it was met with a lot of rejection. Let me say it has not been implemented. So, there are areas of policy that are in conflict with integration and all that should be done away with and there is a need to provide guidance to different counties on how integration should be carried out without being too prescriptive.” (Female respondent, policy maker, T1)

> “I would say it hasn’t been, although it is in policy documents, as in NCDs as a collection of conditions, but the concept of multimorbidity hasn’t been highlighted as much as it should I would say.” (Male respondent, clinician and health economist, T5)

##### Donor priorities (vertical programs)

Additionally, donor-driven funding was considered a challenge to integrated care implementation whereby donors often prioritise specific diseases like HIV, TB, and malaria, limiting focus on integrated care and multimorbidity. This donor influence was considered to perpetuate siloed healthcare delivery, undermining efforts to prioritise and address the rising burden of NCDs comprehensively, as shown in the following remarks from three stakeholders:

> “…So again, due to lack of adequate resources at the ministry our interventions are largely donor driven. With that comes, the conditions of how these funds should be used and, for some it may be very specific to one disease and may not allow for those funds to be used in such a way that integration will be possible, which limits integration” (Female respondent, policy maker, T1)

> “I will say that the heaviest resources from donors go to essential things like vaccination. They go into HIV, TB, malaria and that’s the way it has been, and when we are speaking of traditional donors we are speaking of the government donors and developing partners…priorities in health have heavily been of communicable disease, we’ve not yet seen an active shift to NCDs despite the numbers going up.” (Female respondent, clinician and public health specialist, T12)

> “So, we still have that problem because we still have the funders dictating as much as the country seems to be moving in one direction, I feel that the funders of the main disease conditions are still staying put with siloed care. And that is going to be a problem.” (Female respondent, clinician and public health specialist, T8)

##### Limited funding or budget allocation

Key stakeholders in the study highlighted that limited funding and budget allocation significantly impede the implementation of integrated care for multimorbidity. Despite being recognized as a priority in the NCD strategic plan, efforts have largely remained at the pilot stage due to insufficient financial resources. This lack of funding hampers the development and implementation of necessary guidelines and policies, and the supply and management of commodities at healthcare facilities. The highlighted that while some programs benefit from donor research funding, government investment in integrated care remains inadequate, underscoring a critical gap in financial commitment towards addressing multimorbidity.

> “It has been mainly pilot after pilot if I can say and where have not we and were the ones developing the policy. Why haven’t we developed these guidelines or policies? As we have just said we have lacked funds despite being a priority in the NCD strategic plan” (Female respondent, policy maker, T1)

> “Number two is on the resources, both financial and human. Especially on the financial resources, it directly affects even the commodity supply and managing commodities at those facilities.” (Male respondent, clinician and public health specialist, T13)

> “And I believe the one for Organization X is mainly because it is supported through research, and the like. But in terms of the government investing in that integrated care, that is something that they are missing.” (Female respondent, University lecturer and public health specialist, T9)

##### Limited collaboration between stakeholders or sectors

Despite efforts to integrate NCD prevention into different sector-specific plans, stakeholders indicated a lack of coherent messaging on prevention strategies, stemming from the fragmented operations within programs and divisions. Furthermore, stakeholders outside the health sector are identified as crucial yet underutilised in combating NCDs. Additionally, a gap between practitioners and policymakers, particularly at the county level, was evident.

> “Have we optimised the integration? Even within the ministry of health itself? We have many programs, we have many divisions etc. Do we have a coherent message that is going out on prevention? I know we have tried as much as possible to incorporate NCD prevention into other sector plans.” (Female respondent, policy maker, T2)

> “The other gap as I finish on the question is that we need to bring in stakeholders outside health…There are so many stakeholders that need to come on board.” (Male respondent, University professor and public health specialist, T4)

> “I would say there is also to some extent a gap between the practitioners and the policy makers. Even at the county level that is the key implementation cornerstone, I would say there is a gap between the practitioners, especially the senior specialists at the facility level, and the policy makers…I think that there is an opportunity to bring these people closer…” (Male respondent, clinician and health economist, T5)

### Organisational level

#### Fragmented healthcare delivery

At the organisational level, stakeholders described issues with disease-specific clinics requiring patients to attend multiple appointments for different conditions, leading to inefficiencies and gaps in care. They noted that even current integration efforts often remain siloed, treating conditions like HIV, hypertension, diabetes, and osteoarthritis separately. The need to break down these silos was emphasised, with a call for scalable, low-cost models of integrated care to better address the complex needs of individuals with multiple morbidities as highlighted in the following three quotes.

> “… The obvious one is hypertension clinic, there is a diabetes clinic, there is a clinic for mental health…you find there is a diabetologist in the hospital who is not looking into whether the same patient has hypertension or not. So, integration, I think, is not being really practised very well in our setting.” (Male respondent, clinician and public health specialist, T15)

> “…but I think the policy environment is actually siloed. And how diseases are managed. I know these are pushed towards more integrated care now and person-centred care. But I still see siloing in these…which we probably need to get rid of so that we see the individual as an individual who has multiple needs. and not a single condition.” (Male respondent, clinician and public health specialist, T15)

> “So, we need to ensure that we break the silos. People are doing different things…but we are working in silos, therefore our efforts are not very impactful because they are fragmented. So, there is a need to work together so that we can address these multiple morbidities to ensure impact.” (Male respondent, University professor and public health specialist, T4)

### Functional level

#### Limited resources or commodities to support comprehensive healthcare delivery

Stakeholders involved in clinical work identified significant challenges with drug availability, highlighting frequent stockouts as a major barrier to comprehensive patient care as multiple diseases require more resources including medications. Clinics often face disruptions due to unreliable drug supplies, attributable to inefficiencies at the County level. This inconsistency forces patients to seek care at higher-level hospitals, affecting patient retention. One clinician highlighted:

> “The barriers actually that I can talk about is the lack of drugs, you know we diagnose patients, we bring patients on board but there are no drugs in the facilities. So, you are setting up a clinic, a clinic breaks down because this month there are drugs, the following month there are no drugs. And you know that is the challenge with the county…with any kind of government. The drugs are there for a short period of time then there are stock outs.” (Male respondent, clinician, T7)

The same stakeholder went on to reflect on this challenge at county level in the following quote:

> “…the first thing is to improve drug care at the lower-level facilities. The counties must accept that it is not easy to stock drugs up to the lowest level, they must embrace the revolving fund pharmacy. Even if it is getting the drugs from the higher facilities pushing them down because if a patient keeps coming to the county hospital because of drugs, it causes a lot of challenges…” (Male respondent, clinician, T7)

### Professional level

#### Lack of knowledge or capacity on integrated care delivery

Stakeholders highlighted the critical need to address gaps in knowledge and capacity among healthcare workers as a barrier to effective care integration. Both clinicians and public health specialists emphasised the importance of providing proper training, tools, and awareness to better screen and manage multimorbidity, recognizing these as essential steps for successful upscaling of integrated care services. This was summarised by two stakeholders who highlighted as follows:

> “The last aspect is capacity. Do we have the equipment? Do we have empowered healthcare workers in terms of knowledge and skills? So, providing the right equipment and also empowering these healthcare workers to provide these (integrated) services is one of the key aspects we need to focus on as we plan to upscale.” (Male respondent, clinician and public health specialist, T13)

> “…you need proper sensitisation of healthcare providers to raise awareness about multimorbidity and to raise awareness about screening for multimorbidity.” (Male respondent, clinician and public health specialist, T15)

#### Limited human resources

Participants also highlighted limited human resources as a key barrier to healthcare integration. They emphasised the critical shortage of essential staff, including nurses and physicians, which strains the delivery of integrated services and health education. Addressing this staffing gap is crucial for improving integrated management of chronic diseases. Two stakeholders remarked as follows:

> “The third thing, probably, would be making available staff, adequate staff to provide those (integrated) services or health education that’s required to be given to the general population.” (Male respondent, clinician and health economist, T5)

> “…and also, the number of staff. Again, there are very few nurses and physicians. For example, the county at that time only had one physician who was the main consultant to all these patients who are sick. So, you see he is overworking. The medical officers are few, the clinical officers are few, every cadre that are involved in health - even the health record officers - were very few. The challenge there is staffing.” (Male respondent, clinician, T7)

### Normative integration

#### ‘Siloed’ mindset

Another challenge that emerged was a ‘siloed mindset’ in the management of chronic diseases. It was indicated that different stakeholders operated independently with minimal coordination between them. This fragmentation hinders the effectiveness of efforts to address complex health conditions in an integrated way. One participant highlighted the need to dismantle these silos and promote collaborative strategies to develop scalable integrated care models of care that improve the impact of interventions for multimorbidity, as highlighted in the following two quotes:

> “…it is like people sometimes are working in silos. Yes, community health workers with their things, public health officers, people in the hospitals have their own things and managers again with the budgeting process having their own things sometimes.” (Male respondent, clinician and health economist, T5)

> “So, we need to ensure that we break the silos. People are doing different things. We are working in silos therefore our efforts are not very impactful because they are fragmented. There is a need to work together so that we can address these multiple morbidities or disease challenges to ensure impact. Also, we need to come up with a model which can be scaled” (Male respondent, clinician and health economist, T5)

#### Poor advocacy

Poor advocacy was considered a challenge to implementing integrated care. Stakeholders highlighted the lack of sustained lobbying efforts for integrated management of cardiometabolic diseases in health facilities in the counties. One participant noted that despite healthcare workers’ efforts at the community level, the absence of continuous advocacy limited long-term improvements.

> “…lack of the people to lobby. Let us see when we go to the ground, we do our best but then we are not there throughout sadly enough because cardiovascular care is lacking in the public hospitals you find in many counties.” (Female respondent, clinician and public health specialist, T11)

#### Limited evidence

Stakeholders highlighted gaps in research on cost-effectiveness and the integration of care for communicable and non-communicable diseases. Despite some attempts to integrate hypertension and HIV care, these initiatives have not resulted in concrete policies due to insufficient data on their effectiveness, cost-effectiveness, and sustainability. Participants pointed to the lack of studies on cost-effectiveness, emphasising the need for more economic evaluations to inform decision-making and optimise resource allocation for scaling interventions.

> “There have been a few hypertension-HIV studies that are mainly implemented in the western region, but this has really not translated to tangible policies because there is still a lot of unknown when it comes to integration with communicable diseases.” (Female respondent, policy maker, T1)

> “We have not had a lot of studies around cost-effectiveness of interventions, and I think this is something that we really need to embrace. I think it should be important before we roll out programs or after two years, three years of roll out…it is important to evaluate whether they are cost-effective so that we can invest in the best interventions.” (Female respondent, University lecturer and public health specialist, T9)

### Recommendations for implementation of integrated care for cardiometabolic multimorbidity

#### Systems level

##### Clear and standardised policies or guidelines

The key stakeholders emphasised the need for clear, standardised policies and guidelines to support the implementation and evaluation of integrated care for cardiometabolic diseases in Kenya. They recommended the development of comprehensive, adaptable guidelines that could be applied across various regions while ensuring consistent integration of services, particularly for cardiovascular disease, diabetes, and mental health care. They also stressed the importance of developing practical toolkits for healthcare providers to enhance diagnosis and management of multiple conditions, thereby improving overall patient outcomes. Three stakeholders made the following remarks:

> “I know there was a task sharing policy (an integration concept), but it was met with a lot of rejection. So, there are areas of policy that are in conflict with integration and all that should be done away with, and then there is need to provide a clear guidance in terms of how integration should be carried out in different counties without being too prescriptive.“ (Female respondent, policy maker, T1)

> “But at a higher level, I think we need proper guidelines. I don’t think there are any guidelines that talk about multimorbidity. Right now, we find there are hypertension treatment guidelines, there are diabetes treatment guidelines…but the question is do we pick up people who have multimorbidity as they come for care?” (Male respondent, clinician and public health specialist, T15)

> “I think the first thing, before you even scale up, we need guidelines. Something that will guide the counties and facilities on how they can do integration, provide different options of course, as you see counties are different, and what works in one may not necessarily work in the other…and you need a framework to monitor the extent of integration.” (Female respondent, policy maker, T2)

##### Multiple stakeholder involvement

Most stakeholders interviewed emphasised the importance of involving different stakeholders including community members, healthcare professionals, and political leaders to prioritise and advocate for integrated care. Stakeholders also stressed the need for collaboration among professional bodies to develop comprehensive integrated care guidelines for multimorbidity, covering cardiovascular, diabetes, mental health, and HIV care. They also recommended practical toolkits for healthcare providers to facilitate diagnosis, management, and referral, ensuring better comprehensive healthcare delivery. This is highlighted in the following two quotes:

> “As I said, the same patients you find might be the same ones having these things (chronic diseases). So, until we then also as communities and as the people receiving these services ensure that we also prioritise integrated care. We need voices from the communities, we need the political class voices to push the realisation of tangible integration.” (Female respondent, clinician and public health specialist, T12)

> “…it should be with the role of NCD Inter-sectoral Coordinating Committee (ICC) to find a way of bringing these professional bodies or representatives together and say can we come up with a core multimorbidity care guideline that has key aspects of cardiovascular disease…, key aspects of diabetes, of mental health, of HIV?.” (Female respondent, clinician and public health specialist, T8)

##### Planning and budgeting for integration

Stakeholders emphasised the role of public participation in influencing county-level fiscal planning and budgeting for non-communicable diseases. By involving community members and healthcare workers in fiscal planning cycles, integrated care can be prioritised within county budgets. Furthermore, participants stressed that sustainable financing is critical for multimorbidity management, as a lack of deliberate funding can undermine integration efforts. Structured financial commitments are essential to ensure comprehensive care delivery.

> “So integrated work planning which ensures that everyone is in the room with NCD folks when budgets are being done. Actually, from that county, there is a fiscal cycle - there is something called public participation, and this is across the board. I have seen public participation change priorities for counties, even the care givers or even the healthcare workers at community level ensuring that it is prioritised within the fiscal planning cycle that can really help with that integration.” (Female respondent, clinician and public health specialist, T12)

> “And of course the third one is financing. There has to be deliberate financing in that multimorbidity management and maybe that is at the integrated care level in a way, because of course everything falls apart when we get to the financing bit.” (Female respondent, clinician and public health specialist, T8)

### Professional level

#### Adequately trained staff to support integration

At the professional level, ensuring an adequate number of health care providers at the primary care level trained in integrated management of cardiometabolic diseases (such as integrated cardiovascular disease and diabetes management) with sufficient knowledge, expertise and shared vision of integrated care was recommended by the participants to bridge the limited resources and capacity gap for integrated care. Sensitisation and capacity building of health personnel from the systems to facility level was considered integral to achieving this goal.

This theme is exemplified by the following quotes:

> “…We need to find a way to address that so that the mentors are probably family physicians. So, we can have a family physician in every county, that would be the person who is able to receive mentorship from the specialist and then be the one to mentor the general practitioner rather than direct mentorship from the specialist.” (Female respondent, clinician and public health specialist, T8)

> “…also making sure there is the third thing, probably, would be making available staff, adequate staff to provide those services like health education that are required for the general population.” (Male respondent, clinician and health economist, T5)

> “…you need proper sensitisation of healthcare providers to raise awareness about multimorbidity and to raise awareness about screening for multimorbidity.” (Male respondent, clinician and public health specialist, T15)

### Functional level

#### Resources and systems to support integration

The participants considered resources and systems integral to the delivery of integrated care and therefore should be provided to enable integrated care to be realised. Resources include adequate provision of medications, diagnostics and equipment for screening and management of cardiometabolic diseases including hypertension and diabetes. One participant involved in an integrated care intervention suggested the concept of ‘revolving fund pharmacies’ as a viable solution to drug stock-outs in lower-level facilities.

> “…making available necessary resources such as the medicines, the diagnostics, and equipment,” (Male respondent, clinician and health economist, T5)

> “…to improve drug care at the lower-level facilities, the counties must accept that it is not easy to stock drugs up to the lowest level. They must embrace the revolving fund pharmacies.” (Male respondent, clinician, T7)

### Clinical level

#### Designing integrated healthcare delivery models

A key recommendation was to deliberately design the health system to provide integrated care to manage multimorbidity at the point of health care delivery. It was also acknowledged that reorganising care would be a risk to the quality of care given the challenges of activities such as task shifting, and therefore should be conducted carefully to ensure that the quality of care is not compromised in any way.

> “…there should be deliberate effort to structure programs to address multimorbidity. That is at the health facility level, ensuring that scheduling of patients, aa…capacity of healthcare workers, even allocation of resources in the clinics is commensurate with the burden of multimorbidity” (Male respondent, clinician and public health specialist, T15)

> “It is really a risk we must watch against, because even as we are proposing integration, we must be very careful not to push things. It goes hand in hand with task shifting for you to have integrated care. We must be very careful not to push a cadre of staff to offer care that they are not able to offer.” (Male respondent, clinician and public health specialist, T15)

#### Need for evidence and advocacy

Stakeholders emphasised the need to prioritise research on cost-effective integrated care interventions targeted at high-burden contexts, which they perceived was the role of researchers. They also acknowledged their role in supporting research. Additionally, advocacy was identified as essential for securing leadership buy-in at both national and county levels. Engaging leadership is critical for mobilising the necessary infrastructure and resources to support the integration model of care, ensuring that implementation is effective and sustainable.

> “NCDs are many, money is not there but as a starting point, we need to have interventions that are cost effective and will reach enough people also where the burden is highest, so we need research and prioritisation.” (Female respondent, policy maker, T1)

> “…some of the key aspects that personally I feel, we really need to take up on them and actualise this integration model of care is on the advocacy aspect. We need to bring in the leadership because once the leadership gets a buy-in into this then it becomes easy for us to implement. So, we need to really enhance advocacy, both at the national and the county levels by bringing in the leadership to champion this advocacy then they will champion this model of care…” (Male respondent, clinician and public health specialist, T13)

## Discussion

This study explored stakeholders’ contextual understanding of integrated care, barriers to and recommendations on its implementation at primary care to address cardiometabolic multimorbidity in Kenya. Integrated care was contextually defined by stakeholders mainly from the clinical (holistic care provision), systems (guidelines and standards promoting comprehensive healthcare delivery), and professional (interdisciplinary coordination centred around the healthcare providers) dimensions of the RMIC. Key challenges identified included disparity between policy and practice, inadequate resources, donor-driven priorities, limited stakeholder collaboration, limited capacity and human resources, siloed mindset, poor advocacy and limited evidence. To facilitate effective implementation of integrated care at primary care level, stakeholders recommended developing clear guidelines, enhancing stakeholder engagement, adequate planning and budgeting, improving staff availability and capacity, adequate resource allocation, improving advocacy, evidence, and deliberate configuration of the health system to facilitate integrated care implementation.

The stakeholders defined integrated care as holistic and patient-centred “one-stop shop” delivery of healthcare, and coordination between healthcare providers to deliver health services at primary care level is consistent with the existing integrated care principles and interventions. Interventions such as integrated care clinics have been explored in HICs and are increasingly adopted in LMICs as a ‘one-stop shop” model of service provision [49–51]. While there has been increasing evidence in SSA on the integration of NCDs within HIV services, evidence on integrating NCD services within primary care is scarce [27,52–55]. In Kenya, integration has progressed more successfully for HIV and NCD services [28,53,56] and the integration of maternal and child health services at primary care [57,58], highlighting a foundation upon which further NCD integration efforts could draw lessons upon.

The perceived importance of the professional integration in the Kenyan context aligns with existing literature, which highlights the importance of multidisciplinary teams in delivering comprehensive care for multimorbidity [59–61]. A critical barrier to professional integration in Kenya is the resource limitations within devolved county healthcare systems, where health worker-to-patient ratios fall below WHO recommendations [62]. This shortfall highlights the urgent need for workforce strengthening to support a broader range of chronic conditions, including cardiometabolic diseases. As recommended by stakeholders in this study, training general practitioners or nurses to manage NCDs at PHC level could improve integration outcomes and enable primary care facilities to address the complex needs of patients with multimorbidity, advancing the government’s goal of universal health coverage (UHC). For instance, evidence shows that nurse-led care through task shifting could be effective in improving hypertension and diabetes care at primary care [63–65].

While organisational integration is a key dimension of integrated care [32], it was not considered by stakeholders as a key aspect in their definition of integrated care. This may be explained by the stakeholders’ focus on immediate clinical and professional integration challenges, such as direct patient care and interprofessional collaboration, rather than broader organisational structures, which are often seen as abstract, resource-intensive, or outside their control [59,61]. The findings highlight the need for better organisational integration such as strengthening referral systems between primary care and other levels of care which has been cited as a key challenge in recent studies in Kenya [37].

At the systems level, stakeholders noted a disconnect between policy and practice in regard to integrated care. This finding is consistent with previous studies in LMICs, where policy implementation often fails due to systemic challenges, including limited capacity, resource constraints, and poor inter-sectoral collaboration [23,53,66,67]. Donor-driven funding also emerged as a challenge that complicated integration of care, with resources often earmarked for vertical programs targeting specific diseases such as HIV, TB, and malaria. Evidence from LMICs including Kenya suggests that donor priorities often misalign with local health needs, undermining comprehensive healthcare delivery, highlighting the need to integrate vertical programs into PHC [57,66,68–70]. This integration is most urgent in LMICs transitioning away from donor funding.

The challenge of fragmented healthcare delivery linked to ‘siloed’ mindset in the management of cardiometabolic diseases including cardiovascular diseases and diabetes, imposes significant challenges for patients and the health system. For the patients, fragmented care often means attending separate appointments for hypertension and diabetes on different days, leading to increased out-of-pocket expenses, loss of income due to time away from work, and higher chances of missed appointments, which further compromises their health outcomes. For instance, studies in SSA have shown that the majority of patients with hypertension or diabetes do not know that they have other complications [71,72]. At the healthcare system level, this fragmentation strains already limited resources, particularly at PHC level, where the majority of patients in Kenya seek care, exacerbating inefficiencies and diminishing the capacity to deliver comprehensive and integrated care [23].

The integration of cardiometabolic diseases at PHC level in Kenya requires a multifaceted approach, with several key recommendations emerging from stakeholders. The development of clear, standardised integrated care guidelines tailored to multimorbidity is crucial to address gaps in policy implementation. While the national strategic plan includes the integration of NCDs into existing ministry of health programs [39], there are no specific guidelines on how the integration should be conducted or monitored. This highlights a need to make the guidelines operational and adaptable to the county level, given the diversity of county health systems. This should be accompanied by comprehensive training and advocacy at national and county level to enhance healthcare workers’ capacity, organisational readiness, and culture for integrated care. A study in South Africa that demonstrated the feasibility of implementing an integrated chronic disease management model revealed the benefit of developing user-friendly public sector integrated care guidelines for primary care [73]. In addition to the suggested guidelines, Kenya could use relevant regional guidelines such as WHO African regional committee targets for integrating essential NCD services for the early detection and management of CVD, diabetes, chronic respiratory diseases and cancer in PHC to adapt national and develop county-specific guidelines for the integration of NCD services into PHC [74,75].

The prioritisation of integrated care in national and county-level fiscal plans in Kenya requires evidence on the effectiveness and cost-effectiveness of existing integrated care models. While some integrated care models have recently been piloted in Kenya [51,76,77], there is limited evidence on their effectiveness or long-term cost-effectiveness, highlighting a need for more evidence generation on the effectiveness and cost-effectiveness of different integrated care models. Given the resource constraints that often shape budget allocations in LMICs including Kenya, fiscal policy decisions for health demand for rigorous long-term cost-effectiveness analyses of different models to ensure sustainable investment in integrated care that maximises health outcomes within limited budgets. A recent systematic review found only one study from Kenya assessed the cost-effectiveness of integrated care using a model-based economic evaluation [78], highlighting the need for more evidence.

Integrating vertical programs into the broader health systems framework in Kenya can be achieved by aligning donor funding with integrated care goals. This could be achieved through pooled funding models and partnerships between the government and private programs [68]. Stakeholder engagement is vital, emphasising the need for collaboration between community members, healthcare professionals, researchers, and policy makers in order to secure buy-in at macro-, meso-, and micro-level and foster policy alignment. Lastly, resource allocation for capacity building through training is necessary to equip healthcare workers with the skills and tools required for managing multimorbidity, in addition to ensuring that essential medicines and equipment are consistently available for continuity of care. Resource constraints have been highlighted in different settings in SSA, including Kenya, as a key barrier to the provision of comprehensive services and hence should be addressed to realise effective integrated care delivery [23,53,79].

### Strengths, limitations, and health policy implications

Our study’s strength lies in its qualitative approach, capturing a wide range of stakeholder perspectives, including policy-level, non-governmental policy or advocacy, research, academia, clinicians, and a health economist, which provided a nuanced understanding of the challenges and recommendations for integrating cardiometabolic diseases at PHC in Kenya. However, the limited representation of stakeholders at the county level and non-inclusion of patients in this study is a potential limitation. Based on the purposeful selection of the key stakeholders, our findings from the interviews may not be generalisable to all stakeholders nationally. However, the stakeholders’ perspectives provide valuable insights that can guide health policy and practice for NCD prevention and control. The findings highlight the need for multi-stakeholder engagement including policy makers, researchers including health economists, county health managers, and healthcare providers in measuring, evaluating, and adapting cost-effective context-specific integrated care models tailored to population needs. The findings enable researchers including health economists to formulate and test hypotheses for different integrated care models to furnish policy makers the crucial evidence needed to influence practice.

## Conclusion

The findings underscore the need for policy and practice reforms to promote the integration of cardiovascular disease and diabetes at PHC level in Kenya. Stakeholders at national and county levels should establish clear, standardised guidelines adaptable at county level, enhance resource allocation for multimorbidity management, expand workforce capacity through training, align donor policies, and foster collaboration. Supporting evidence generation of integrated care through funding of effectiveness and cost-effectiveness studies will promote evidence-based prioritisation and resource allocation for integrated care. This will provide valuable insights into how integrated care can be adapted, scaled, and sustained to meet the unique needs of Kenya’s health system and effectively manage the growing burden of multimorbidity.

## Data Availability

Data generated or analysed during this study are available upon request from the following co-authors: Elvis Wambiya -, James Oguta -, Pete Dodd -.

## Acknowledgements

The authors acknowledge the study participants for their valuable time and contributions for this study. We also acknowledge two research assistants, Mr. Solomon Toweet and Mr. Romeo Ngesa of Moi University for supporting the project with verbatim transcription of the audio files.

## Author information

### Contributions

EW, JO, RA, DG, and PD contributed to the conceptualization, study design, and methodology for this study. EW and JO contributed to data collection. EW, JO, RA, DG, PD, and CA contributed to formal data analysis. EW wrote the original draft of the manuscript, and JO, RA, DG, PK, PO, CA, JK, YK, GG, and PD supported review and editing of the manuscript. Supervision was provided by RA, DG, and PD. All authors approved the final version of the manuscript.

## Funding

Funding support for this study was provided by Wellcome as part of a doctoral training grant [218462/Z/19/Z] at the University of Sheffield.

## Ethics declarations Competing interests

The authors declare no competing interests.

## Notes

### Competing Interest Statement

The authors have declared no competing interest.

